# Efficacy and Safety of Middle Meningeal Artery Embolization for Patients with Chronic Subdural Hematoma: A Systematic Review and Meta-Analysis

**DOI:** 10.1101/2024.07.23.24310607

**Authors:** N Kabir, B Owais, G Trifan, FD Testai

## Abstract

**Background:** Chronic subdural hematoma (CSDH) is characterized by the collection of blood beneath the dura mater. Traditional treatments involve surgical drainage of the hematoma, but recurrence rates can be high. A highly vascularized neo-membrane irrigated by the middle meningeal artery (MMA) may be involved in CSDH re-accumulation.

**Objective:** We conducted a systematic review and meta-analysis of studies that compared the efficacy and safety of MMA embolization to conventional treatment alone for CSDH.

**Material and Methods:** A systematic search of PubMed, Embase Ovid, and ClinicalTrials.gov identified observational and randomized clinical studies comparing MMA embolization to conventional treatment for chronic subdural hematoma. The efficacy outcomes were hematoma recurrence and good functional outcome (as defined by a modified Rankin Scale score (mRS) of 0-2). Safety outcomes were the rate of major complication and mortality. Heterogeneity among studies were evaluated using the I^2^ statistic. Analyses were conducted using Cochrane Review Manager software, with risk ratios (RR) and 95% confidence intervals (95% CI) presented for key outcomes. Absolute risk reduction (ARR, 95% CI) 1000 patients were also calculated using GRADEpro software.

**Results:** The analysis included data from 13 studies (4 RCTs and 9 observational studies) with a total number of 2960 patients (35.3% in the MMA group and 64.7% in the conventional treatment group). Compared to conventional treatment, MMA embolization decreased risk of hematoma recurrence by 60% (13 studies, RR=0.40, 95% CI 0.25-0.63; I^2^=50%), for an absolute effect of 119 fewer events/1000 patients (95% CI 70-149), with similar risk of major complications (9 studies, RR=0.82, 95% CI=0.54-1.25) and mortality risk (13 studies, RR=0.90, 95% CI=0.54-1.51). In subgroup analyses by study type, pooled results from RCTs showed similar direction effects as those from observational studies for both efficacy and safety outcomes.

**Conclusion:** MMA embolization in CSDH management is a safe and effective approach for CSDH.

## Introduction

Chronic subdural hematoma (CSDH) refers to the accumulation of blood, blood-degradation products, of fluid between the dura mater and the arachnoid. CSDH affects preponderantly the elderly and has an estimated incidence of 1.7 to 20.6 per 100,000 persons per year.^1^ However, based on the increase in global life expectancy and the trends obtained in observational studies, it is projected that the incidence of this condition will continue to rise^2^. The traditional management of CSDH, which includes surgical drainage, is associated with periprocedural complications and recurrence rates that range from 3% to 30%^3^. Historically, it was considered that CSDH was the consequence of venous oozing secondary to traumatic injury of the bridging veins. However, cumulative evidence indicates that, although trauma, even if mild, may constitute an initial insult, the pathogenesis of CSDH involves the development of a layer of connective tissue cells that leads to the formation of a new membrane^4^. This membrane is enriched in pro-inflammatory and pro-angiogenic mediators, such as tissue-plasminogen activators, matrix-metalloproteinases, and vascular endothelial growth factor. These mediators contribute to the development of leaky vessels that result in repeated microbleeds and the exudation of fluid. This phenomenon further enhances inflammation and results in a vicious cycle that perpetuates CSDH and contributes to its recurrence.

Since the vascular supply to this new membrane derives from the middle meningeal artery (MMA), it has been proposed that the embolization of the MMA may reduce the risk of hematoma growth and recurrence. Since the MMA is a branch of the external carotid artery, it was considered that this procedure would not significantly increase the risk of cerebrovascular complications. Data supporting the benefit and safety of endovascular embolization of the MMA as an adjunct to surgical drainage originates from non-randomized retrospective studies and registries. In a meta-analysis including non-randomized studies, MMA embolization had lower rates of CSDH recurrence and need for surgical rescue with comparable rates of in-hospital complications^5^. The results of three randomized clinical trials (RCTs) assessing the benefits and risks of MMA embolization as an adjunct therapy for patients with CSDH were recently reported^6,7,8,9–11^. Here we present the results of a meta-analysis where we assessed the efficacy and safety of MMA for the treatment of patients with CSDH.

## Material and Methods

Our study follows the Preferred Reporting Items for Systematic Reviews and Meta-Analyses (PRISMA) statement guidelines^12^. This protocol is registered in PROSPERO, trial number IDCRD42024515860. Data is available from the corresponding author upon reasonable request. No informed consent was required from the human subjects given the nature of the analysis.

### Eligibility Criteria and Outcomes

This study included randomized controlled clinical trials and observational studies that compared the efficacy and safety of MMA embolization plus standard of care (conservative or surgical treatment) to standard of care alone for patients with CSDH. Studies that only reported outcomes for MMA embolization without separately reporting the outcomes for conventional management only, case reports, and non-peer-reviewed studies were excluded. The primary efficacy outcome was the risk of hematoma recurrence, and the safety outcome was mortality risk. In a secondary analysis, we investigated the risk of good functional outcomes defined as modified Rankin Scale Score (mRS) of 0-2 at follow-up and the frequency of major complications. **Supplemental Table 1** depicts the definitions used in each trial. Given the wildly variable definitions for the safety endpoints used in the RCTs (**Supplementary Table 2**), we only report here the rates of safety points from each RCT, while presenting the pooled results of safety outcomes of the observational studies.

### Information Sources and Search Methods

We queried PubMed, Embase Ovid, and ClinicalTrials.gov from inception to February 2024. The search algorithm is shown in **Appendix 1**. The results were screened for inclusion criteria by 2 of the authors (GT and FDT). Disagreements were resolved using the other two authors (NK and BO). Information on trial design, sample size, mean age, sex, laterality of the hematoma (unilateral vs bilateral), etiology, baseline CSDH volume or thickness, and mean follow-up time, along with the safety and efficacy outcomes were collected.

### Statistical Analyses and Bias Assessment

Two of the authors (GT and FDT) independently determined the risk of bias in RCTs using the RoB2 (Revised Cochrane Risk of Bias) tool^13^; the risk of bias in nonrandomized studies was assessed using the intervention (ROBINS-I) tool^14^. Publication bias was investigated by visual inspection of funnel plots for asymmetry for each reported outcome. Heterogeneity between studies was quantified using *I*^2^ statistic, and as recommended by the American Heart Association^15^, using the following cut-offs: 0–40%: might not be important; 30–60%: may represent moderate heterogeneity; 50–90%: may represent substantial heterogeneity; and 75– 100%: considerable heterogeneity. To further investigate heterogeneity, subgroup analysis by study design was performed.

Analyses were done using Cochrane Review Manager (v 5.4)^16^. Descriptive characteristics of the included studies were recorded. Event rates for each efficacy and safety outcome of interest were compared between MMA embolization plus conventional treatment and conventional management alone. Results are presented as risk ratios along with their corresponding 95% confidence intervals (CI), using random-effects Mantel-Haenszel method. The absolute number of events per 1000 patients (absolute risk reduction, 95% CI) was calculated using the GRADEpro software^17^.

## Results

### Study Selection and Characteristics

The search yielded a total of 3,978 records. Of these, 13 studies with 2,960 patients (35.3% in the MMA group and 64.7% in the conventional treatment group) were included in the final analysis (**Figure 1**). There were 9 observational studies and 4 RCTs. Three of the RCTs were recently presented at the 2024 International Stroke Conference^6,7,8,9–11^. Baseline characteristics of the included studies are presented in **Table 1**. In the MMA group, mean (SD) age was 72.4±4.6 years, 73.3% were males, and mean SDH thickness was 18± 4.6. Comparatively, in the MMA group mean age was 70.6± 5.4 years, 70% were males, and the mean SDH thickness was 18.5± 4.7 mm. Mean follow-up time between groups was similar among groups (3.2 months). The risk of bias across all studies, and for each included study is shown in **Supplementary Figure 1**. Visual inspection of the funnel plots revealed symmetry for the main outcomes (**Supplementary Figure 2**).

**Figure 1.**
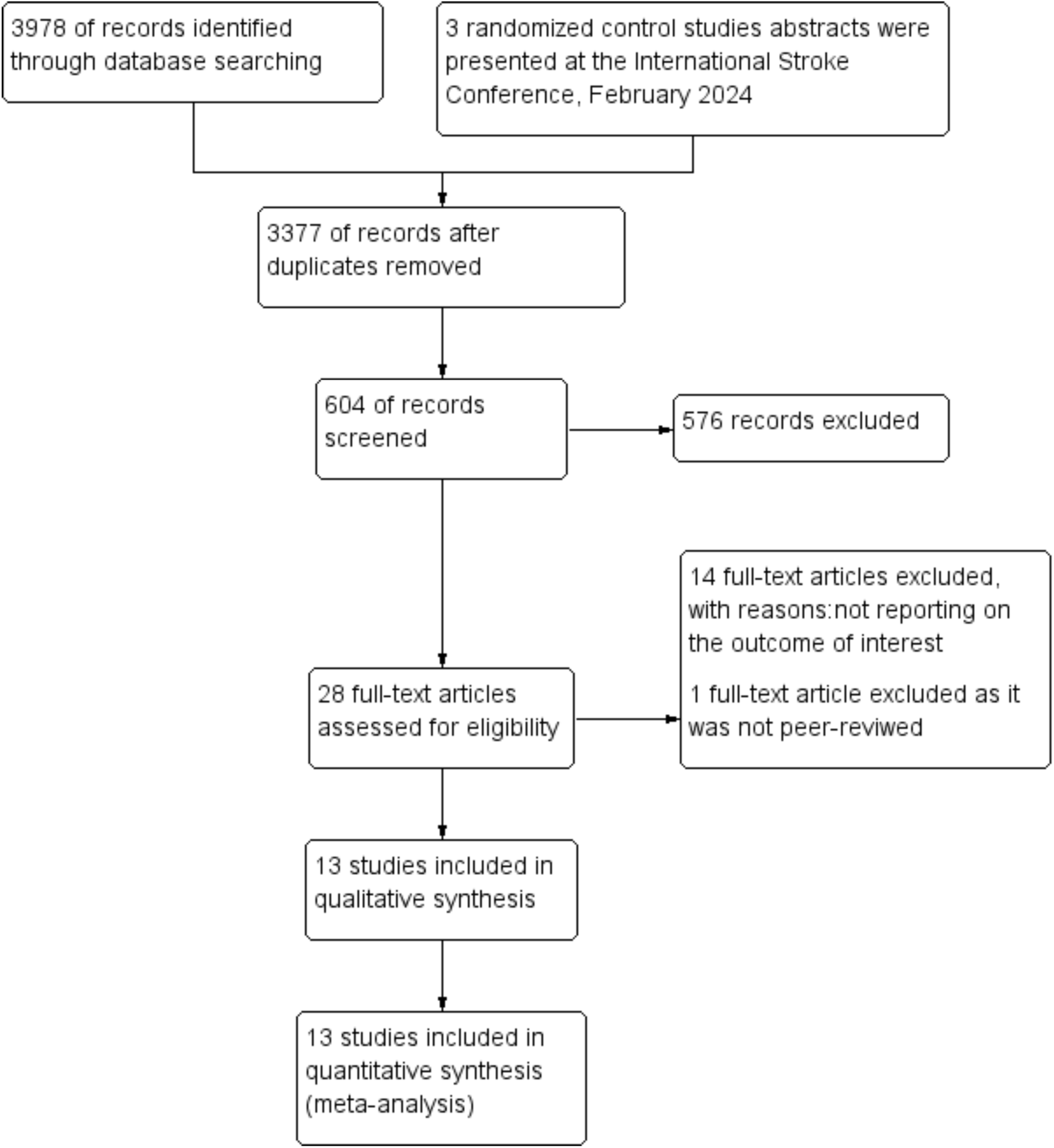
PRISMA Flow Diagram.

**Table 1.**
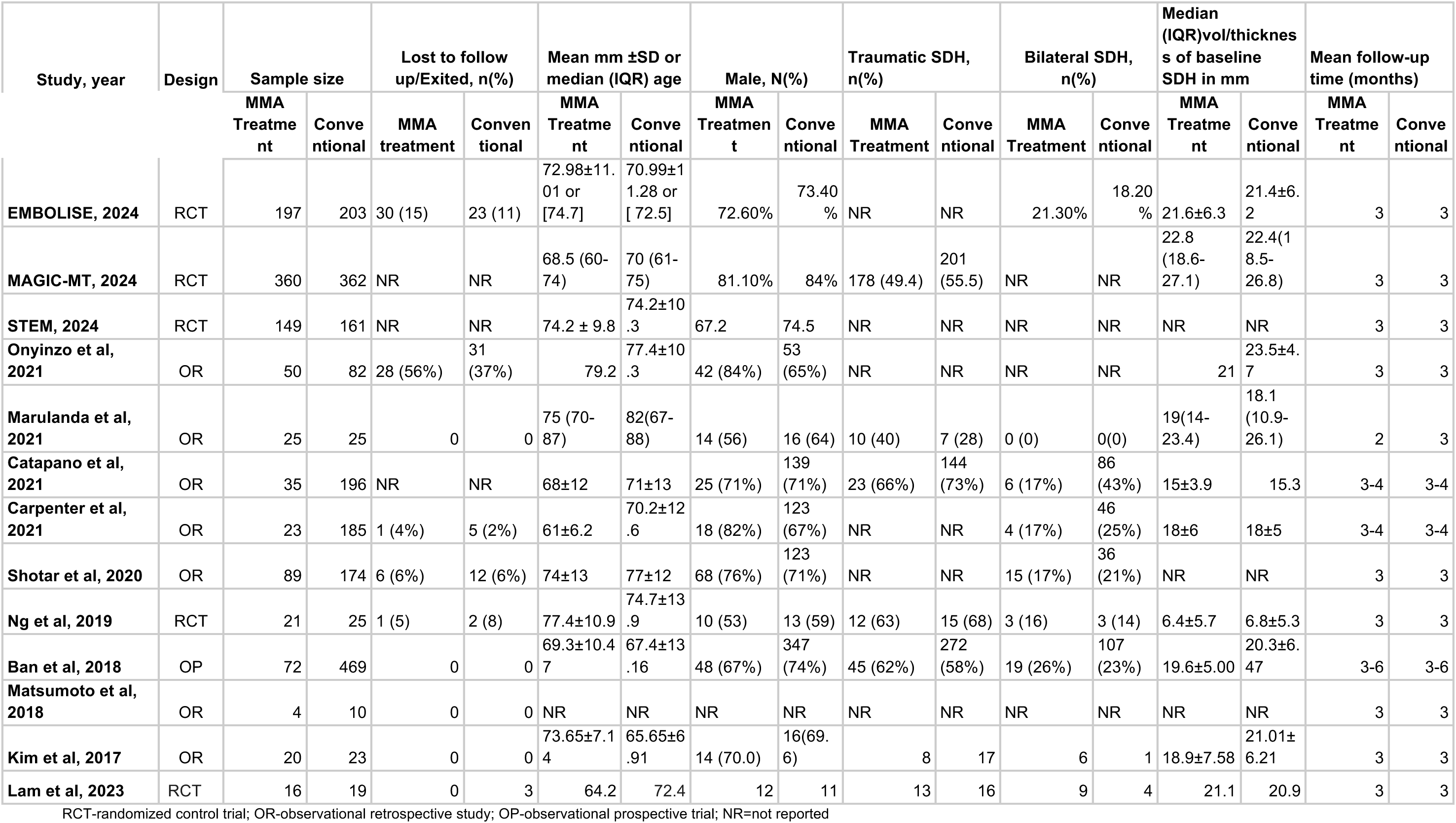
Baseline Characteristics^5^.

### Efficacy outcomes

**Figure 2** depicts the risk of hematoma recurrence between adjunct MMA embolization and conventional management alone for all studies. There was an absolute effect of 119 fewer events per 1,000 patients (95% CI 70-149) and an overall risk reduction of 60% in favor of adjunct MMA embolization compared with conventional treatment alone (13 studies, RR=0.40, 95% CI 0.25-0.63) with moderate heterogeneity (I^2^=50%). When analyzing by study design, data from RCTs showed a risk reduction of hematoma recurrence with adjunct MMA embolization of 52% (4 studies, RR=0.48, 95% CI=0.34-0.67), with low heterogeneity (I^2^=15%) and an absolute effect of 86 fewer hematoma recurrences per 1,000 participants (95% CI 54-109) (**Figure 2**). In comparison, observational studies showed a 74% risk reduction for hematoma recurrence in favor of adjunct MMA (RR=0.26, 95% CI= 0.10-0.66) with substantial heterogeneity (I^2^=64%) and an absolute effect of 167 fewer recurrences per 1,000 participants (95% CI 63-203) (**Figure 2**).

**Figure 2.**
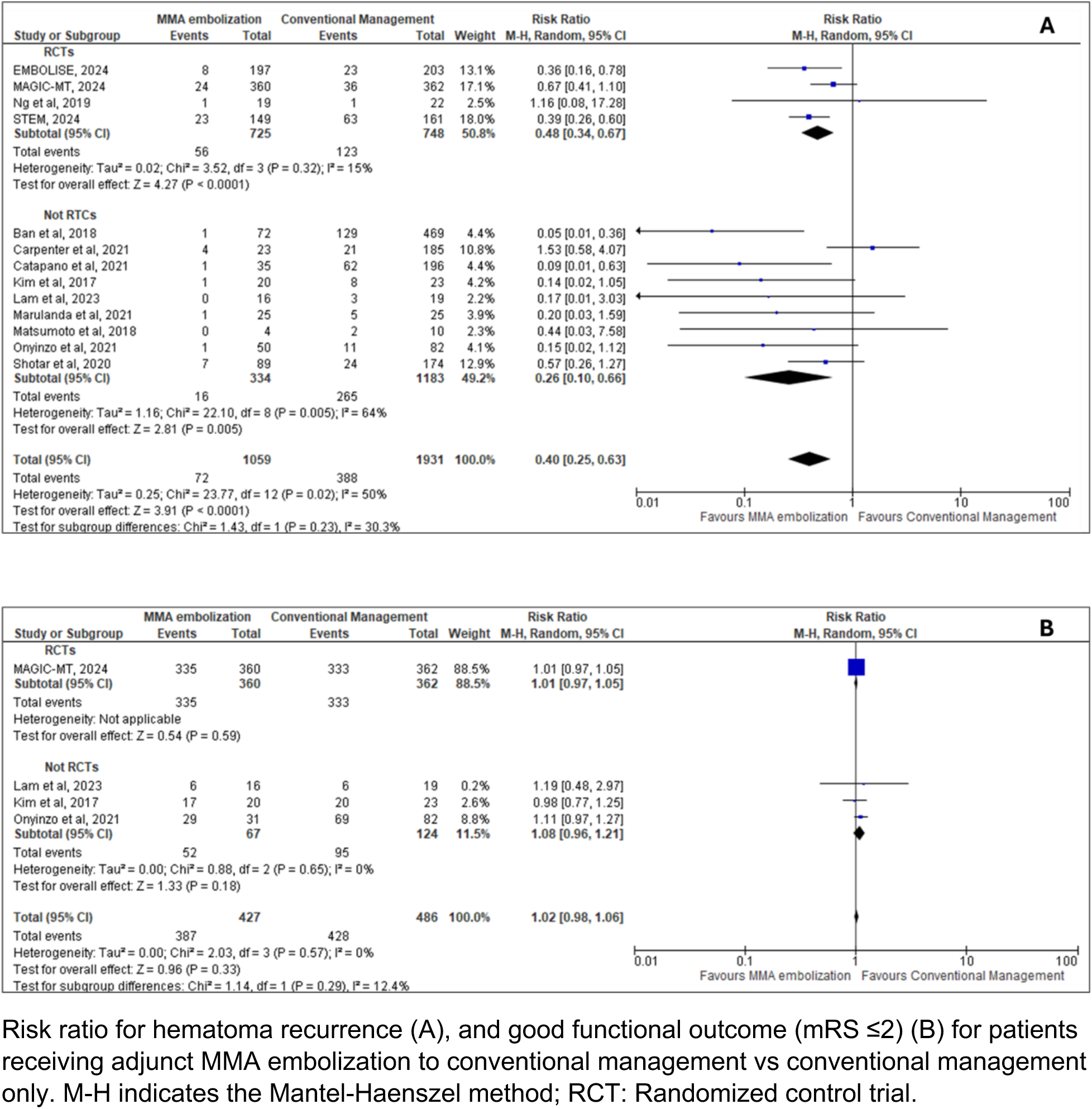
Efficacy outcome with and without MMA embolization.

Rates for good functional outcomes were available for 4 studies only (1 RCT and 3 observational studies). Overall risk for good functional outcome was similar between groups (RR=1.02, 95% CI=0.98 -1.06) with no heterogeneity (I^2^=0%) (**Figure 2**).

### Safety outcomes

**Figure 3** depicts the risk of major complications and mortality rates. Overall, the pooled risk of major complications for all observational studies was 29% lower in the MMA group compared with conventional treatment alone (9 studies, RR=0.82, 95% CI 0.54-1.25) with no heterogeneity (I^2^=0%). The safety endpoint rates reported in each RCT varied between 5% to 16% for the MMA group, compared to 4% to 22% in the conventional treatment group. Mortality rates were similar as well for both groups (13 studies, RR=0.90, 95% CI=0.54 -1.51) with no heterogeneity (I^2^=0%). When analyzed by study design, mortality rates were comparable for RCTs (RR=0.82, 95% CI 0.28-2.34; I^2^=45%) and observational studies (RR=0.87, 95% CI=0.42-1.81, I^2^=0%) (**Figure 3**).

**Figure 3.**
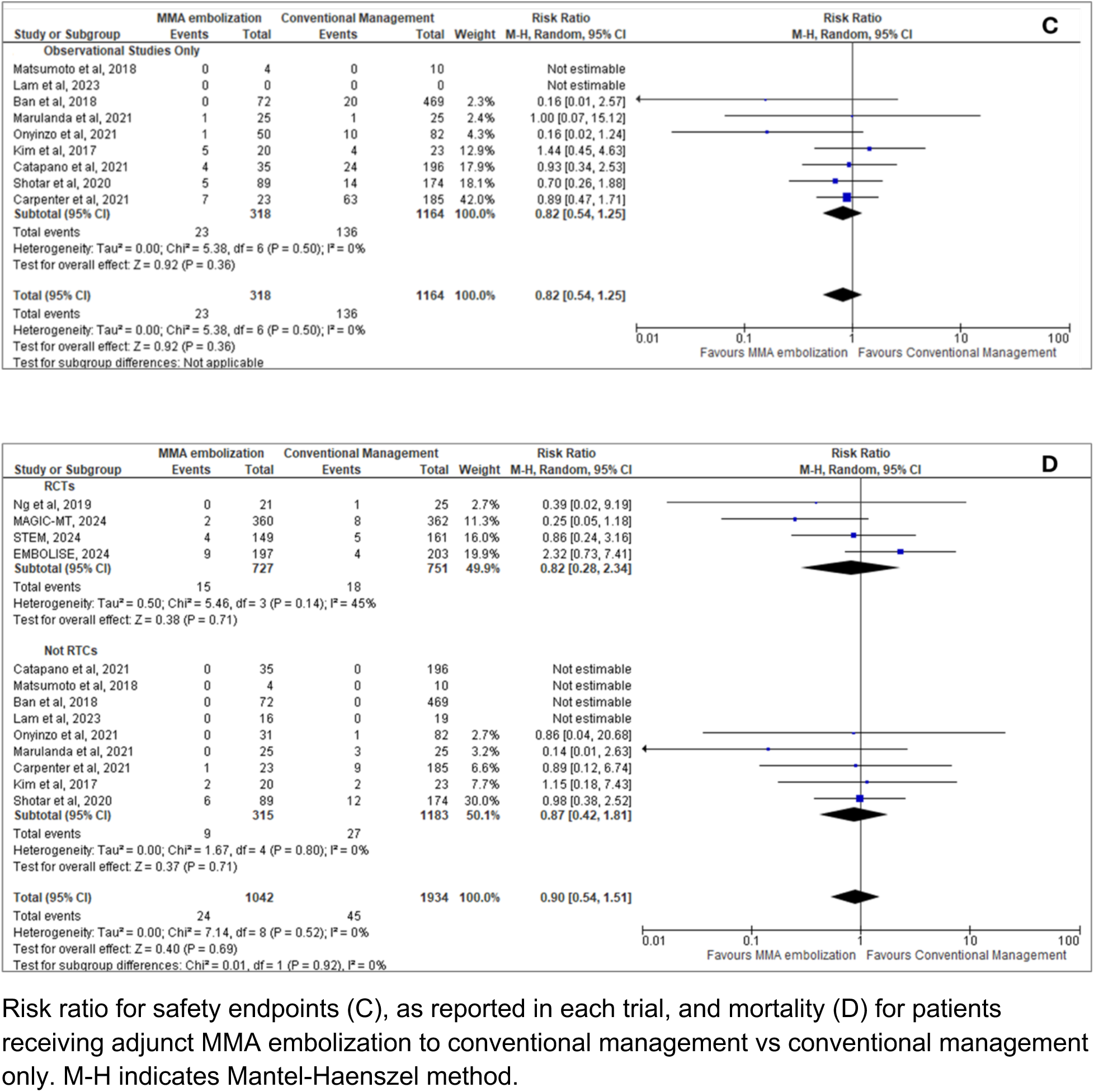
Safety Outcomes with and without MMA embolization.

## Discussion

The results of our meta-analysis show that MMA embolization added to conventional treatment is superior to conventional treatment alone for the treatment of CSDH. Previous meta-analyses that included data from retrospective cohorts have shown that MMA embolization is associated with lower rates of hematoma recurrence and fewer complications^15^. However, the lack of data from RCTs confounded the interpretation of these results. Limitations like retrospective data and lack of separate reporting for conventional treatment hampered their generalizability^5^. This updated meta-analysis addresses these issues by including recent RCTs and observational studies with direct comparisons between MMA embolization with adjunct standard care and standard care alone. In our study, which included the results from three new RCTs^6,7,8^, we observed a 60% reduction in overall hematoma recurrence risk in favor of MMA embolization plus conventional treatment compared to conventional treatment alone, for an absolute risk reduction of 86 fewer hematoma recurrences per 1000 participants. In the subgroup analysis by study design, the rate of hematoma recurrence reduction in favor of MMA was higher in observational studies than in RCTs (74% vs. 52%). These differences could be explained, at least in part, by methodologic factors, including patient selection, publication bias, and study design. Also, several studies reported outcomes obtained in small cohorts which can increase the type II error. Furthermore, the wide confidence intervals and moderate heterogeneity observed in observational studies suggest elevated variability in the reported outcomes and raise concerns about the precision of the calculated point estimates. In comparison, the analysis using data from RCTs produced estimates with narrower confidence intervals and low heterogeneity. Thus, the inclusion of adequately powered RCTs allows for a more accurate estimation of the effect of MMA embolization on CSDH outcomes. This finding strengthens the evidence for the efficacy of MMA embolization in chronic subdural hematoma.

The surgical interventions used for the treatment of CSDH, like burr-hole evacuation, are considered safe. However, the rate of perioperative complications and mortality observed in CSDH cohorts varies from 0% to 32%. These complications are not always caused by the surgical procedure itself but are secondary to other factors such as preexisting active medical conditions and frailty^18,19^. MMA embolization, which is typically done endovascularly, is well tolerated and is also considered safe. The MMA contributes minimally to cerebral blood flow, and collateral circulation can effectively compensate for the blocked MMA, minimizing the risk of cerebrovascular complications. Additionally, embolization is performed via the external carotid artery, further reducing the risk of ischemic stroke^20^. The overall incidence of complications in cohorts treated with MMA embolization is less than 4%^21^. The safety of MMA embolization is also supported by the results of previous meta-analyses which reported similar complication rates for adjunct MMA embolization and conventional treatment groups. ^5^ ^22^ In our study, there were no significant differences in the safety endpoint or mortality for the pooled results, indicating that MMA embolization plus conventional treatment provides a safe alternative to conventional management alone for CSDH. Future studies should assess the long-term durability of the MMA effect and whether this approach allows the use of anticoagulation in patients who require such treatment. Similar to previous studies, we observed that MMA embolization as an adjunct treatment does not improve functional outcomes compared to conventional management alone^22^. However, it should be noted that most of the studies included in this analysis ascertained functional outcomes using the mRS. This scale is widely used in stroke research. However, the sensitivity of this tool to detect differences in disability in the particular case of SDH, where the neurologic deficits may be subtle, has not been well described.

### Limitations

Our meta-analysis has several limitations. First, the conventional treatment was not standardized among studies; thus, there are intrinsic variations among the studies that we are not able to account for. Second, there is variability in the definitions of efficacy and safety outcomes among studies. Third, the majority of the studies included in our analysis had an observational retrospective design with the consequent risk of selection bias^23^. In addition, many of them were performed in small cohorts and did not include power calculations. However, in subgroup analysis, we observed that the results reported in observational studies are comparable to those obtained in adequately powered RCTs. Fourth, we reported our results as RRs and absolute effects. Because the absolute risk for control varies between trials, these estimates should be interpreted with caution. However, our meta-analysis is the largest study comparing the efficacy and safety of MMA as adjunct therapy to standard treatment alone for patients with CSDH. In addition, we included the results of three large RCTs which, combined, accounted for almost 75% of the final sample size and allowed us to calculate point estimates with low heterogeneity.

## Conclusion

MMA embolization as an adjunct treatment significantly decreases the recurrence of CSDH without increasing the risk of morbidity or mortality when compared to conventional treatment alone. Several studies are currently underway to compare the efficacy and safety of MMA embolization as a standalone treatment for CSDH.

## Data Availability

All data referred in the manuscript has reference associated with it.

## Supplemental Data

**Supplementary Figure 1:**
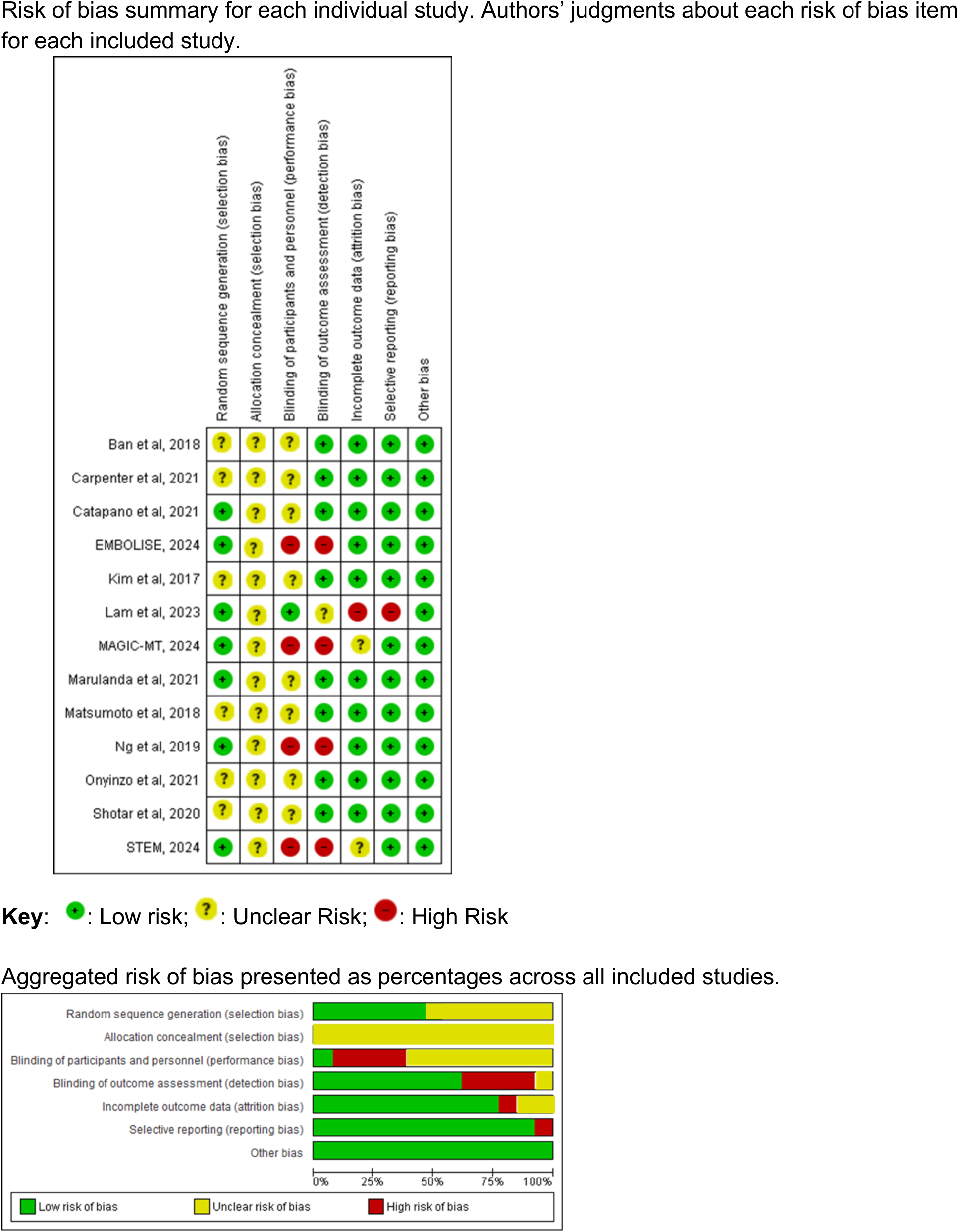
Risk of bias across and between studies.

**Supplementary Figure 2:**
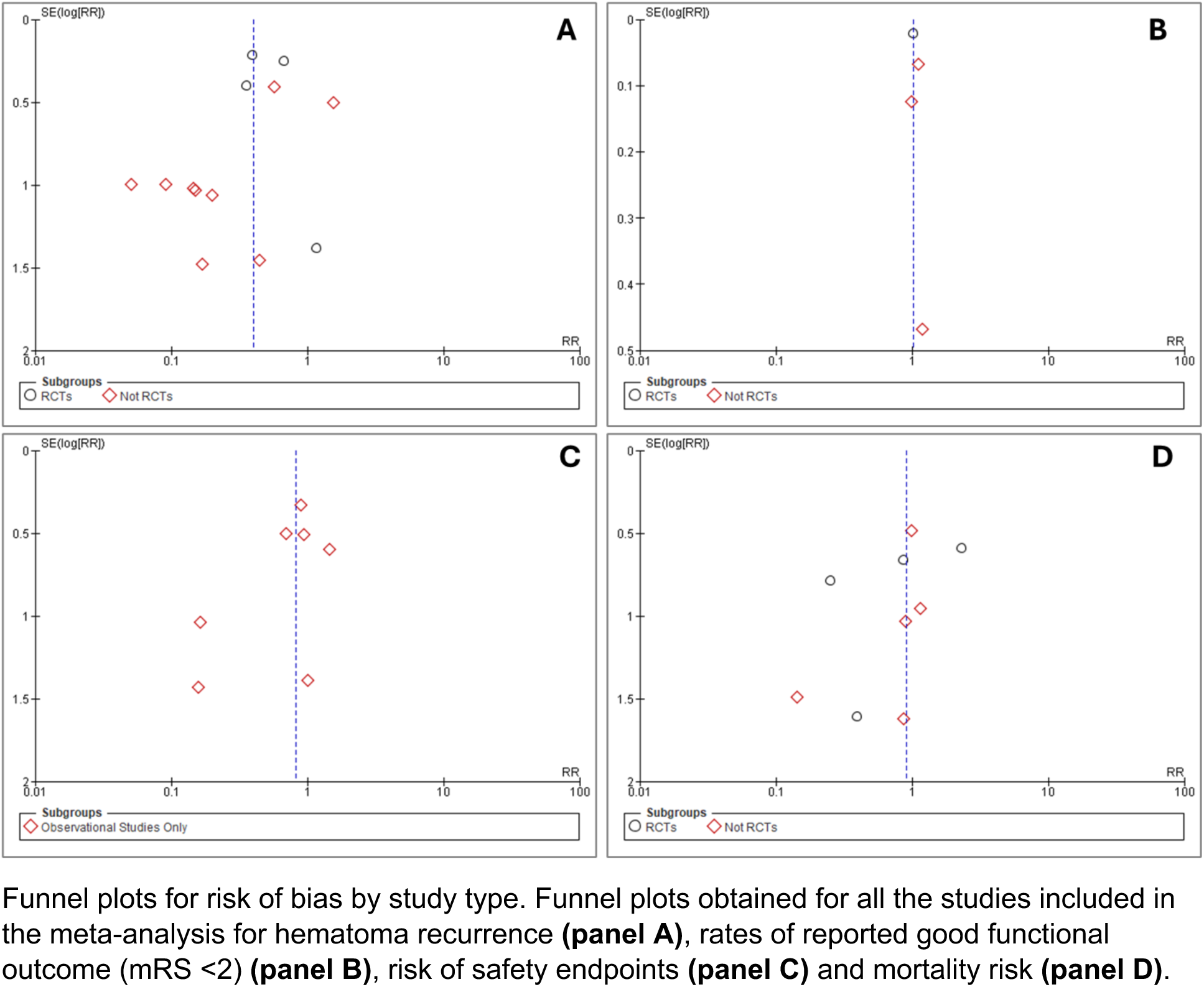
Risk of bias plots.

**Supplemental Table 1.** Hematoma Recurrence as defined in each study.

| Study | Hematoma Recurrence |
| --- | --- |
| EMBOLISE, 2024 | hematoma recurrence/progression requiring surgical drainage |
| MAGIC-MT, 2024 | hematoma maximum thickness exceeding 10 mm at 90 days or the patient receiving re-operation within 90 days in patients who underwent burr-hole drainage |
| STEM, 2024 | Residual or re-accumulation of the SDH ($\geq 10$ mm) on 180-day scan from intervention |
| Onyinzo et al, 2021 | CT findings in the follow-up period residual CSDH > 10mm |
| Marulanda et al, 2021 | Recurrence was defined as reaccumulating hematoma or continuous growth discovered on follow-up imaging with or without a new acute/subacute component |
| Catapano et al, 2021 | residual cSDH or reaccumulation > 10 mm |
| Carpenter et al, 2021 | re-accumulation of hematoma with symptoms requiring reoperation |
| Shotar et al, 2020 | reoperation to treat a recurrent CSDH previously treated by burr-hole craniostomy |
| Ng et al, 2019 | any radiologically persistent subdural collection with persistent or new symptoms attributable to CSDH |
| Ban et al, 2018 | incomplete hematoma resolution (remaining or reaccumulated hematoma with thickness > 10 mm |
| Matsumoto et al, 2018 | residual hematoma thickness of more than 50% or the absence of size reduction |
| Kim et al, 2017 | increase in the hematoma density and width |
| Salih et al, 2023 | SDH expansion after intervention or appearance of new acute components that warranted intervention or continuous close clinical and radiographic follow-up |
| Lam et al, 2023 | radiologically persistent or new cSDH with persistent or new symptoms secondary to the mass effect of the cSDH |

**Supplemental Table 2.**
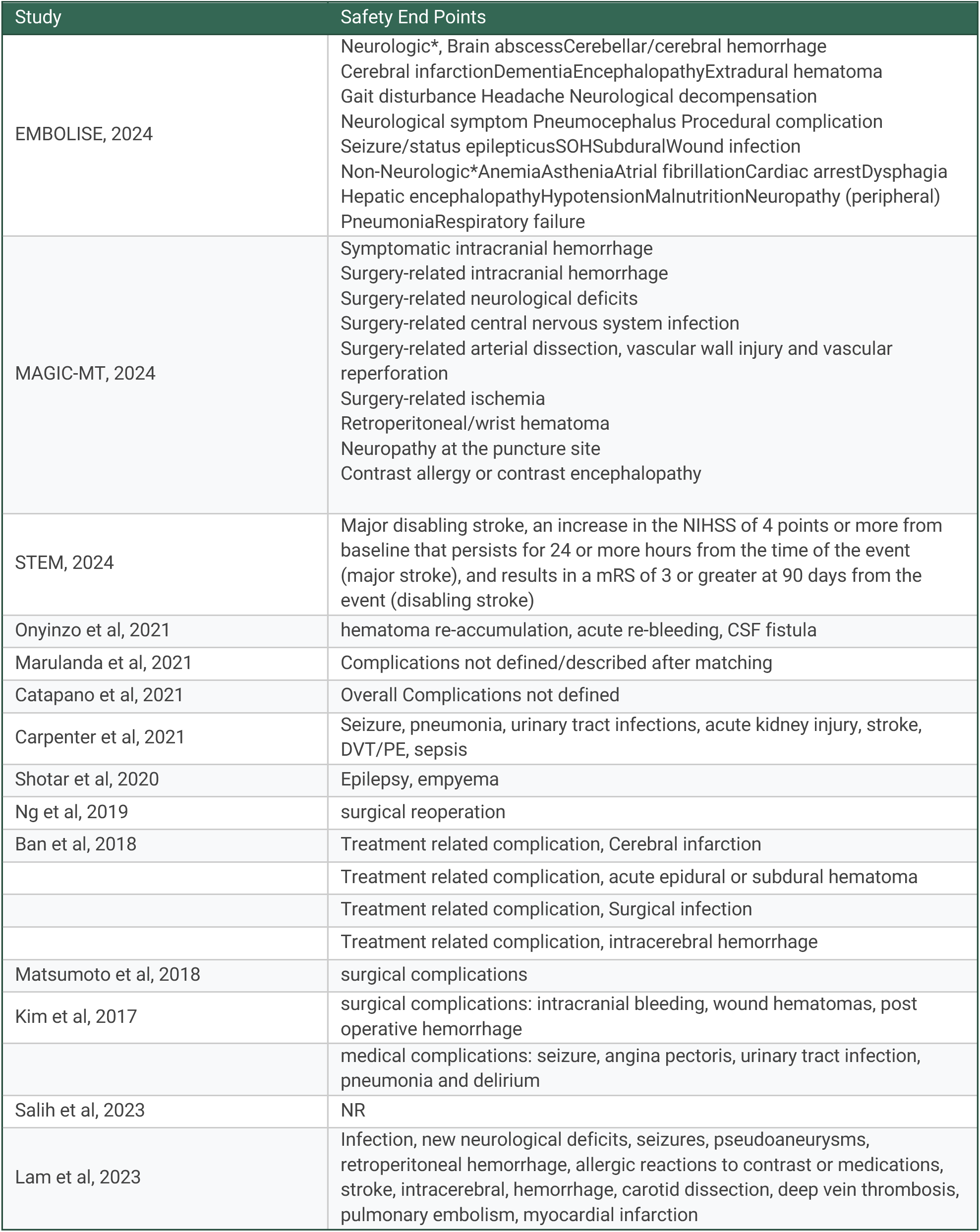
Safety End Points as defined in each study.

## Appendix 1 Embase

### Search Algorithm

……………………………………………….

No. Query Results

#5. (((’subdural hematoma’/exp OR ’subdural hematoma’) AND middle AND meningeal AND artery AND embolization OR chronic) AND subdural AND hemorrhage OR hematoma) AND recurrence AND embolization

#4. (((’subdural hematoma’/exp OR ’subdural hematoma’) AND middle AND meningeal AND artery AND embolization OR chronic) AND subdural AND hemorrhage OR hematoma) AND recurrence AND embolization

#3. ((’subdural hematoma’/exp OR ’subdural hematoma’) AND middle AND meningeal AND artery AND embolization OR chronic) AND subdural AND hemorrhage

#2. (((’subdural hematoma’/exp OR ’subdural hematoma’) AND middle AND meningeal AND artery AND embolization OR chronic) AND subdural AND hemorrhage OR hematoma) AND recurrence

#1. ’subdural hematoma’/exp OR ’subdural hematoma’

……………………………………………….

Results 601

601

1,653

9,574

24,688

### Pubmed search algorithm

Search number Query Sort By Filters Search Details Results

4 (((((Middle Meningeal Artery Embolization) AND (chronic subdural hematoma)) AND (outcomes)) AND (surgery)) OR (burr hole)) AND (recurrence) ((("middle"[All Fields] OR "middles"[All Fields]) AND ("meningeal arteries"[MeSH Terms] OR ("meningeal"[All Fields] AND "arteries"[All Fields]) OR "meningeal arteries"[All Fields] OR ("meningeal"[All Fields] AND "artery"[All Fields]) OR "meningeal artery"[All Fields]) AND ("embol"[All Fields] OR "embolics"[All Fields] OR "embolisations"[All Fields] OR "embolise"[All Fields] OR "embolised"[All Fields] OR "embolising"[All Fields] OR "embolism"[MeSH Terms] OR "embolism"[All Fields] OR "embolic"[All Fields] OR "embolisms"[All Fields] OR "embolization, therapeutic"[MeSH Terms] OR ("embolization"[All Fields] AND "therapeutic"[All Fields]) OR "therapeutic embolization"[All Fields] OR "embolisation"[All Fields] OR "embolization"[All Fields] OR "embolizations"[All Fields] OR "embolize"[All Fields] OR "embolized"[All Fields] OR "embolizes"[All Fields] OR "embolizing"[All Fields]) AND ("chronic subdural haematoma"[All Fields] OR "hematoma, subdural, chronic"[MeSH Terms] OR ("hematoma"[All Fields] AND "subdural"[All Fields] AND "chronic"[All Fields]) OR "chronic subdural hematoma"[All Fields] OR ("chronic"[All Fields] AND "subdural"[All Fields] AND "hematoma"[All Fields])) AND ("outcome"[All Fields] OR "outcomes"[All Fields]) AND ("surgery"[MeSH Subheading] OR "surgery"[All Fields] OR "surgical procedures, operative"[MeSH Terms] OR ("surgical"[All Fields] AND "procedures"[All Fields] AND "operative"[All Fields]) OR "operative surgical procedures"[All Fields] OR "general surgery"[MeSH Terms] OR ("general"[All Fields] AND "surgery"[All Fields]) OR "general surgery"[All Fields] OR "surgery s"[All Fields] OR "surgerys"[All Fields] OR "surgeries"[All Fields])) OR ("trephining"[MeSH Terms] OR "trephining"[All Fields] OR ("burr"[All Fields] AND "hole"[All Fields]) OR "burr hole"[All Fields])) AND ("recurrance"[All Fields] OR "recurrence"[MeSH Terms] OR "recurrence"[All Fields] OR "recurrences"[All Fields] OR "recurrencies"[All Fields] OR "recurrency"[All Fields] OR "recurrent"[All Fields] OR "recurrently"[All Fields] OR "recurrents"[All Fields]) 710

3 ((((Middle Meningeal Artery Embolization) AND (chronic subdural hematoma)) AND (outcomes)) AND (surgery)) OR (burr hole) (("middle"[All Fields] OR "middles"[All Fields]) AND ("meningeal arteries"[MeSH Terms] OR ("meningeal"[All Fields] AND "arteries"[All Fields]) OR "meningeal arteries"[All Fields] OR ("meningeal"[All Fields] AND "artery"[All Fields]) OR "meningeal artery"[All Fields]) AND ("embol"[All Fields] OR "embolics"[All Fields] OR "embolisations"[All Fields] OR "embolise"[All Fields] OR "embolised"[All Fields] OR "embolising"[All Fields] OR "embolism"[MeSH Terms] OR "embolism"[All Fields] OR "embolic"[All Fields] OR "embolisms"[All Fields] OR "embolization, therapeutic"[MeSH Terms] OR ("embolization"[All Fields] AND "therapeutic"[All Fields]) OR "therapeutic embolization"[All Fields] OR "embolisation"[All Fields] OR "embolization"[All Fields] OR "embolizations"[All Fields] OR "embolize"[All Fields] OR "embolized"[All Fields] OR "embolizes"[All Fields] OR "embolizing"[All Fields]) AND ("chronic subdural haematoma"[All Fields] OR "hematoma, subdural, chronic"[MeSH Terms] OR ("hematoma"[All Fields] AND "subdural"[All Fields] AND "chronic"[All Fields]) OR "chronic subdural hematoma"[All Fields] OR ("chronic"[All Fields] AND "subdural"[All Fields] AND "hematoma"[All Fields])) AND ("outcome"[All Fields] OR "outcomes"[All Fields]) AND ("surgery"[MeSH Subheading] OR "surgery"[All Fields] OR "surgical procedures, operative"[MeSH Terms] OR ("surgical"[All Fields] AND "procedures"[All Fields] AND "operative"[All Fields]) OR "operative surgical procedures"[All Fields] OR "general surgery"[MeSH Terms] OR ("general"[All Fields] AND "surgery"[All Fields]) OR "general surgery"[All Fields] OR "surgery s"[All Fields] OR "surgerys"[All Fields] OR "surgeries"[All Fields])) OR ("trephining"[MeSH Terms] OR "trephining"[All Fields] OR ("burr"[All Fields] AND "hole"[All Fields]) OR "burr hole"[All Fields]) 3,978

2 (hematoma, subdural, chronic (MeSH)’ OR ‘chronic" OR ‘subdural hematoma’ AND ‘meningeal arteries (MeSH)’ OR ‘middle meningeal artery’ AND ‘embolization, therapeutic’ (MeSH) OR ‘embolization’ AND ‘recurrence’) OR (embolization and recurrence) (((((((((("hematoma, subdural, chronic"[MeSH Terms] OR ("hematoma"[All Fields] AND "subdural"[All Fields] AND "chronic"[All Fields]) OR "chronic subdural hematoma"[All Fields] OR "hematoma subdural chronic"[All Fields]) AND ("medical subject headings"[MeSH Terms] OR ("medical"[All Fields] AND "subject"[All Fields] AND "headings"[All Fields]) OR "medical subject headings"[All Fields] OR "mesh"[All Fields])) OR ("chronic"[All Fields] OR "chronical"[All Fields] OR "chronically"[All Fields] OR "chronicities"[All Fields] OR "chronicity"[All Fields] OR "chronicization"[All Fields] OR "chronics"[All Fields]) OR ("subdural haematoma"[All Fields] OR "hematoma, subdural"[MeSH Terms] OR ("hematoma"[All Fields] AND "subdural"[All Fields]) OR "subdural hematoma"[All Fields] OR ("subdural"[All Fields] AND "hematoma"[All Fields]))) AND ("meningeal arteries"[MeSH Terms] OR ("meningeal"[All Fields] AND "arteries"[All Fields]) OR "meningeal arteries"[All Fields])) AND ("medical subject headings"[MeSH Terms] OR ("medical"[All Fields] AND "subject"[All Fields] AND "headings"[All Fields]) OR "medical subject headings"[All Fields] OR "mesh"[All Fields])) OR (("middle"[All Fields] OR "middles"[All Fields]) AND ("meningeal arteries"[MeSH Terms] OR ("meningeal"[All Fields] AND "arteries"[All Fields]) OR "meningeal arteries"[All Fields] OR ("meningeal"[All Fields] AND "artery"[All Fields]) OR "meningeal artery"[All Fields]))) AND ("embolization, therapeutic"[MeSH Terms] OR ("embolization"[All Fields] AND "therapeutic"[All Fields]) OR "therapeutic embolization"[All Fields] OR "embolization therapeutic"[All Fields])) AND ("medical subject headings"[MeSH Terms] OR ("medical"[All Fields] AND "subject"[All Fields] AND "headings"[All Fields]) OR "medical subject headings"[All Fields] OR "mesh"[All Fields])) OR ("embol"[All Fields] OR "embolics"[All Fields] OR "embolisations"[All Fields] OR "embolise"[All Fields] OR "embolised"[All Fields] OR "embolising"[All Fields] OR "embolism"[MeSH Terms] OR "embolism"[All Fields] OR "embolic"[All Fields] OR "embolisms"[All Fields] OR "embolization, therapeutic"[MeSH Terms] OR ("embolization"[All Fields] AND "therapeutic"[All Fields]) OR "therapeutic embolization"[All Fields] OR "embolisation"[All Fields] OR "embolization"[All Fields] OR "embolizations"[All Fields] OR "embolize"[All Fields] OR "embolized"[All Fields] OR "embolizes"[All Fields] OR "embolizing"[All Fields])) AND ("recurrance"[All Fields] OR "recurrence"[MeSH Terms] OR "recurrence"[All Fields] OR "recurrences"[All Fields] OR "recurrencies"[All Fields] OR "recurrency"[All Fields] OR "recurrent"[All Fields] OR "recurrently"[All Fields] OR "recurrents"[All Fields])) OR (("embol"[All Fields] OR "embolics"[All Fields] OR "embolisations"[All Fields] OR "embolise"[All Fields] OR "embolised"[All Fields] OR "embolising"[All Fields] OR "embolism"[MeSH Terms] OR "embolism"[All Fields] OR "embolic"[All Fields] OR "embolisms"[All Fields] OR "embolization, therapeutic"[MeSH Terms] OR ("embolization"[All Fields] AND "therapeutic"[All Fields]) OR "therapeutic embolization"[All Fields] OR "embolisation"[All Fields] OR "embolization"[All Fields] OR "embolizations"[All Fields] OR "embolize"[All Fields] OR "embolized"[All Fields] OR "embolizes"[All Fields] OR "embolizing"[All Fields]) AND ("recurrance"[All Fields] OR "recurrence"[MeSH Terms] OR "recurrence"[All Fields] OR "recurrences"[All Fields] OR "recurrencies"[All Fields] OR "recurrency"[All Fields] OR "recurrent"[All Fields] OR "recurrently"[All Fields] OR "recurrents"[All Fields])) 19,903

1 (hematoma, subdural, chronic (MeSH)’ OR ‘chronic’ OR ‘subdural hematoma’ AND ‘meningeal arteries (MeSH)’ OR ‘middle meningeal artery’ AND ‘embolization, therapeutic’ (MeSH) OR ‘embolization’ AND ‘recurrence’) OR (conventional management) (((((((((("hematoma, subdural, chronic"[MeSH Terms] OR ("hematoma"[All Fields] AND "subdural"[All Fields] AND "chronic"[All Fields]) OR "chronic subdural hematoma"[All Fields] OR "hematoma subdural chronic"[All Fields]) AND ("medical subject headings"[MeSH Terms] OR ("medical"[All Fields] AND "subject"[All Fields] AND "headings"[All Fields]) OR "medical subject headings"[All Fields] OR "mesh"[All Fields])) OR ("chronic"[All Fields] OR "chronical"[All Fields] OR "chronically"[All Fields] OR "chronicities"[All Fields] OR "chronicity"[All Fields] OR "chronicization"[All Fields] OR "chronics"[All Fields]) OR ("subdural haematoma"[All Fields] OR "hematoma, subdural"[MeSH Terms] OR ("hematoma"[All Fields] AND "subdural"[All Fields]) OR "subdural hematoma"[All Fields] OR ("subdural"[All Fields] AND "hematoma"[All Fields]))) AND ("meningeal arteries"[MeSH Terms] OR ("meningeal"[All Fields] AND "arteries"[All Fields]) OR "meningeal arteries"[All Fields])) AND ("medical subject headings"[MeSH Terms] OR ("medical"[All Fields] AND "subject"[All Fields] AND "headings"[All Fields]) OR "medical subject headings"[All Fields] OR "mesh"[All Fields])) OR (("middle"[All Fields] OR "middles"[All Fields]) AND ("meningeal arteries"[MeSH Terms] OR ("meningeal"[All Fields] AND "arteries"[All Fields]) OR "meningeal arteries"[All Fields] OR ("meningeal"[All Fields] AND "artery"[All Fields]) OR "meningeal artery"[All Fields]))) AND ("embolization, therapeutic"[MeSH Terms] OR ("embolization"[All Fields] AND "therapeutic"[All Fields]) OR "therapeutic embolization"[All Fields] OR "embolization therapeutic"[All Fields])) AND ("medical subject headings"[MeSH Terms] OR ("medical"[All Fields] AND "subject"[All Fields] AND "headings"[All Fields]) OR "medical subject headings"[All Fields] OR "mesh"[All Fields])) OR ("embol"[All Fields] OR "embolics"[All Fields] OR "embolisations"[All Fields] OR "embolise"[All Fields] OR "embolised"[All Fields] OR "embolising"[All Fields] OR "embolism"[MeSH Terms] OR "embolism"[All Fields] OR "embolic"[All Fields] OR "embolisms"[All Fields] OR "embolization, therapeutic"[MeSH Terms] OR ("embolization"[All Fields] AND "therapeutic"[All Fields]) OR "therapeutic embolization"[All Fields] OR "embolisation"[All Fields] OR "embolization"[All Fields] OR "embolizations"[All Fields] OR "embolize"[All Fields] OR "embolized"[All Fields] OR "embolizes"[All Fields] OR "embolizing"[All Fields])) AND ("recurrance"[All Fields] OR "recurrence"[MeSH Terms] OR "recurrence"[All Fields] OR "recurrences"[All Fields] OR "recurrencies"[All Fields] OR "recurrency"[All Fields] OR "recurrent"[All Fields] OR "recurrently"[All Fields] OR "recurrents"[All Fields])) OR (("conventional"[All Fields] OR "conventionals"[All Fields]) AND ("manage"[All Fields] OR "managed"[All Fields] OR "management s"[All Fields] OR "managements"[All Fields] OR "manager"[All Fields] OR "manager s"[All Fields] OR "managers"[All Fields] OR "manages"[All Fields] OR "managing"[All Fields] OR "managment"[All Fields] OR "organization and administration"[MeSH Terms] OR ("organization"[All Fields] AND "administration"[All Fields]) OR "organization and administration"[All Fields] OR "management"[All Fields] OR "disease management"[MeSH Terms] OR ("disease"[All Fields] AND "management"[All Fields]) OR "disease management"[All Fields])) 91,460

ClinicalTrials.gov

NCT Number

Interventions

NCT04410146

for Treatment of Chronic Subdural Hematoma (STEM)

https://clinicaltrials.gov/study/NCT04410146 ACTIVE_NOT_RECRUITING Subdural Hematoma, Chronic DEVICE: SQUID Embolization|DEVICE: SQUID Embolization and Surgical Evacuation|PROCEDURE: Surgical Evacuation|OTHER: Other: Medical Management Balt USA INTERVENTIONAL

Study Title Study URL Study Status Conditions

Sponsor Collaborators Study Type

The SQUID Trial for the Embolization of the Middle Meningeal Artery

NCT03307395 Middle Meningeal Artery Embolization for Treatment of Chronic Subdural Hematoma https://clinicaltrials.gov/study/NCT03307395 COMPLETED Chronic Subdural Hematoma PROCEDURE: Middel Meningeal Artery Embolization Weill Medical College of Cornell University INTERVENTIONAL

NCT04500795 Prospective Study on the Use of Middle Meningeal Artery Embolization for Chronic Subdural Haematoma https://clinicaltrials.gov/study/NCT04500795 WITHDRAWN Chronic Subdural Hematoma|Subdural Hematoma PROCEDURE: Middle meningeal artery embolization Chinese University of Hong Kong

INTERVENTIONAL

NCT04270955 Dartmouth Middle Meningeal Embolization Trial (DaMMET)

https://clinicaltrials.gov/study/NCT04270955 COMPLETED Chronic Subdural Hematoma|Subdural Hematoma PROCEDURE: Embolization of the Middle Meningeal Artery|PROCEDURE: Standard of care including possible surgical evacuation of subdural hematoma Dartmouth-Hitchcock Medical Center

INTERVENTIONAL

NCT04511572 Embolization of Middle Meningeal Artery in Chronic Subdural Hematoma https://clinicaltrials.gov/study/NCT04511572 RECRUITING Chronic Subdural Hematomas PROCEDURE: embolization of the middle meningeal artery Academisch Medisch Centrum - Universiteit van Amsterdam (AMC-UvA) INTERVENTIONAL NCT04574843 Middle Meningeal Artery Embolization With Liquid Embolic Agent for Treatment of Chronic Subdural Hematoma https://clinicaltrials.gov/study/NCT04574843 RECRUITING Chronic Subdural Hematoma PROCEDURE: Embolization of MMA Mashhad University of Medical Sciences

INTERVENTIONAL

NCT05374681 Efficacy of a Minimally Invasive Therapy Adjuvant to the Standards of Care by Cyanoacrylate Embolization https://clinicaltrials.gov/study/NCT05374681 RECRUITING Chronic Subdural Hematoma OTHER: Medical treatment|OTHER: Surgical treatment|OTHER: embolization of the MMA University Hospital, Brest

INTERVENTIONAL

NCT04742920 The Onyxâ„¢ Trial For The Embolization Of The Middle Meningeal Artery For Chronic Subdural Hematoma (OTEMACS) https://clinicaltrials.gov/study/NCT04742920 RECRUITING Hematoma, Subdural, Chronic|Brain Diseases|Central Nervous System Diseases|Wounds and Injuries

DEVICE: Middle Meningeal Artery Embolization|PROCEDURE: Standard Management University Hospital, Montpellier INTERVENTIONAL

NCT04095819 Middle Meningeal Artery (MMA) Embolization Compared to Traditional Surgical Strategies to Treat Chronic Subdural Hematomas (cSDH)

https://clinicaltrials.gov/study/NCT04095819 UNKNOWN Chronic Subdural Hematoma PROCEDURE: Middle Meningeal Artery procedure|PROCEDURE: Traditional Surgery Atlantic Health System INTERVENTIONAL

NCT06274580 Endovascular vs Conservative Treatment in Patients With Chronic Subdural Hematomas and Mild Symptoms https://clinicaltrials.gov/study/NCT06274580

NOT_YET_RECRUITING Subdural Hematoma, Chronic DEVICE: embolization of the middle meningeal artery Ospedale Policlinico San Martino

INTERVENTIONAL

NCT04816591 Middle Meningeal Artery Embolization for the Treatment of Subdural Hematomas With TRUFILLÂ® n-BCA https://clinicaltrials.gov/study/NCT04816591 RECRUITING Chronic Subdural Hematoma DEVICE: Experimental: Interventional Cohort: Treatment Arm|OTHER: Active Comparator: Interventional Cohort: Control Arm|DEVICE: Experimental: Observational Cohort: Treatment Arm|OTHER: Active Comparator: Observational Cohort: Control Arm Cerenovus, Part of DePuy Synthes Products, Inc. INTERVENTIONAL NCT04065113 Middle Meningeal Artery Embolization for Chronic Subdural Hematoma https://clinicaltrials.gov/study/NCT04065113 RECRUITING Chronic Subdural Hematoma PROCEDURE: Middle Meningeal Artery Embolization with polyvinyl alcohol particles (PVA)|PROCEDURE: Drainage of Subdural Hematoma Washington University School of Medicine INTERVENTIONAL

NCT05327933 Preventing Recurrences of Chronic Subdural Hematoma in Adult Patients by Middle Meningeal Artery Embolization https://clinicaltrials.gov/study/NCT05327933 RECRUITING Hematoma, Subdural, Chronic PROCEDURE: Surgery plus endovascular MMA embolization|PROCEDURE: Surgery alone Unfallkrankenhaus Berlin INTERVENTIONAL

NCT06163547 Middle Meningeal Artery Embolization for Chronic Subdural Hematomas (STORMM) https://clinicaltrials.gov/study/NCT06163547 NOT_YET_RECRUITING Chronic Subdural Hematomas|Cerebral Compression Due to Injury RADIATION: MMA embolization University Hospital, Geneva INTERVENTIONAL

NCT04372147 Embolization of the Middle Meningeal Artery for the Prevention of Chronic Subdural Hematoma Recurrence in High Risk Patients (EMPROTECT) https://clinicaltrials.gov/study/NCT04372147 UNKNOWN Chronic Subdural Hematoma|at Risk of Post-operative Recurrence|Burr-hole Surgery PROCEDURE: MMA embolization Assistance Publique - HÃ pitaux de Paris INTERVENTIONAL

